# A landscape of the genetic and cellular heterogeneity in Alzheimer disease

**DOI:** 10.1101/2021.11.30.21267072

**Authors:** Logan Brase, Shih-Feng You, Ricardo D’Oliveira Albanus, Jorge L. Del-Aguila, Yaoyi Dai, Brenna C Novotny, Carolina Soriano-Tarraga, Taitea Dykstra, Maria Victoria Fernandez, John P Budde, Kristy Bergmann, John C Morris, Randall J Bateman, Richard J Perrin, Eric McDade, Chengjie Xiong, Alison Goate, Martin Farlow, Jasmeer P. Chhatwal, Peter Schofield, Helena Chui, Dominantly Inherited Alzheimer Network (DIAN), Greg T Sutherland, Jonathan Kipnis, Celeste M Karch, Bruno A Benitez, Carlos Cruchaga, Oscar Harari

**Affiliations:** Department of Psychiatry, Washington University School of Medicine, St. Louis, MO, USA; Hope Center for Neurological Disorders, Washington University School of Medicine, St. Louis, MO, USA; NeuroGenomics and Informatics, Department of Psychiatry Washington University in St. Louis; Knight Alzheimer Disease Research Center, Washington University School of Medicine, St. Louis, MO, USA; Department of Neurology, Washington University School of Medicine, St. Louis, MO, USA; Department of Pathology and Immunology, Washington University School of Medicine, St. Louis, MO, USA; Center for Brain Immunology and Glia (BIG), Washington University in St. Louis, St. Louis, MO, USA; Department of Genetics, Washington University School of Medicine, St. Louis, MO, USA; School of Medical Sciences and Charles Perkins Centre, Faculty of Medicine and Health, The University of Sydney, Sydney, NSW, Australia; Dominantly Inherited Alzheimer Network (DIAN); Baylor College of Medicine, Houston, TX, USA; Merck & Co., Inc., Boston, MA, USA; Division of Biostatistics, Washington University School of Medicine, St. Louis, MO, USA; Ronald M. Loeb Center for Alzheimer’s Disease, Department of Genetics and Genomic Sciences, Icahn School of Medicine at Mount Sinai, New York, NY, USA; Department of Neurology, Indiana University School of Medicine, Indianapolis, IN, USA; Department of Neurology, Massachusetts General Hospital, Harvard Medical School, Boston, MA, USA; Neuroscience Research Australia, Sydney, NSW, Australia; School of Medical Sciences, University of New South Wales, Sydney, NSW, Australia; Alzheimer’s Disease Research Center, Keck School of Medicine at the University of Southern California, Los Angeles, CA, USA; Department of Neurology, Keck School of Medicine, University of Southern California, Los Angeles, CA, USA

**Author notes:** To whom correspondence should be addressed: Oscar Harari, PhD, Assistant Professor, Department of Psychiatry, Washington University School of Medicine, Campus Box 8134 | 4444 Forest Park Avenue | Office: 5587 | St. Louis, MO 63108, |. These authors contributed equally (Celeste M Karch, Bruno A Benitez, Carlos Cruchaga, Oscar Harari).

**Keywords:** Single-nucleus RNA sequencing, Alzheimer disease, functional genomics, autosomal dominant Alzheimer disease

## Abstract

**Background:** Alzheimer disease (AD) has substantial genetic, molecular, and cellular heterogeneity associated with its etiology. Much of our current understanding of the main AD molecular events associated with the amyloid hypothesis (*APP, PSEN1* and *PSEN2*) and neuroimmune modulation (*TREM2* and *MS4A*) is based on genetic studies including GWAS. However, the functional genes, downstream transcriptional ramifications, and the cell-type-specific effects of many GWAS loci remain poorly understood. Understanding these effects can point us to the cellular processes involved in AD and uncover potential therapeutic targets.

**Methods:** We applied a genetic-based approach to our sample selection; our cohort included carriers of AD pathogenic mutations (*APP, PSEN1)*, risk variants in *TREM2*, and the resilience variant (rs1582763) in the *MS4A* cluster associated with cerebrospinal fluid (CSF) soluble TREM2 levels. We performed single-nucleus RNA-sequencing (snRNA-seq) of 1,102,459 nuclei from the human parietal cortex of these carriers. Following initial unbiased clustering and cell-type annotation, we performed deep subclustering analysis per cell type to identify unique cellular transcriptional states associated with these genetic variants. We identified differentially expressed genes between cell states and genetic variant carriers/controls, and performed differential cell proportion analyses to determine key differences among these carriers. We analyzed sequencing data from human dorsolateral prefrontal cortex and mouse models to replicate the enrichment of unique cell states in genetic variant carriers. Finally, we leveraged these cell-state differential expression results to link genes in AD GWAS loci to their functional cell types.

**Findings:** We identified cell-specific expression states influenced by AD genetic factors for neurons and glia. Autosomal dominant AD (ADAD) brains exhibited unique transcriptional states in all cell types. *TREM2* variant carrier brains were also enriched for specific microglia and oligodendrocyte subpopulations. Carriers of the resilience *MS4A* variant were enriched for an altered activated-microglia expression state. We mapped AD GWAS genes to their potential functional cell types, and some, including *PLCG2* and *SORL1*, were expressed in a broader range of brain cell types than previously reported.

**Interpretation:** AD pathogenic, risk, or resilience variants are sufficient to alter the transcriptional and cellular landscape of human brains. Overall, our results suggest that the genetic architecture contributes to the cortical cellular heterogeneity associated with disease status, which is a critical factor to consider when designing drug trials and selecting the treatment program for AD patients.

Our findings suggest that integrating genetic and single-cell molecular data facilitates our understanding of the heterogeneity of pathways, biological processes and cell types modulated by genetic risk factors for AD.

**Funding:** US National Institutes of Health, Hope Center, Archer foundation, Alzheimer Association, CZI.

## Main

Alzheimer disease (AD) is the most common form of dementia. It is characterized by the presence of amyloid (Aβ) plaques and neurofibrillary tangles (NFTs, Tau deposits) in the brain and associated with neuroinflammation, myelination changes, synaptic dysfunction, and cell death^1,2^. Genetic studies have successfully identified multiple genes and pathways associated with AD etiology including rare mutations in *amyloid precursor protein* (*APP*)^3,4^, *presenilin 1* (*PSEN1*)^5^, and *presenilin 2* (*PSEN2*)^6^ which cause familial autosomal dominant AD (ADAD), mainly due to the neuronal amyloid cascade hypothesis^7^. The strongest risk factor for sporadic AD (sAD), *apolipoprotein E* (*APOE)* ε4, implicates cholesterol metabolism and Aβ clearance mediated by astrocytes and microglia^8^. Integration of epigenetics and genome-wide association studies (GWAS) signals have implicated immune system dysfunction in AD^9^. In particular, rare genetic variants in t*riggering receptor expressed on myeloid cells 2* (*TREM2*), a gene that drives microglia activation, increase AD risk^10^. Recent GWAS have identified additional common genetic risk factors for AD, including rs1582763, an intergenic variant in the *MS4A* locus that confers resilience to AD and is associated with higher cerebrospinal fluid (CSF) soluble TREM2 levels^11-14^. However, linkage and association studies do not reveal the downstream transcriptional ramifications nor the cell types these variants affect.

Single-nucleus RNA-sequencing (snRNA-seq) has emerged as a powerful approach to interrogate the underlying transcriptional landscape of the cellularly-complex human brain. SnRNA-seq studies of sAD and *TREM2* p.R47H variant carriers to date have profiled the dorsolateral prefrontal cortex (DLPFC)^15,16^, entorhinal cortex^17^, and temporal neocortex^17^, uncovering significant transcriptional changes in microglia, astrocytes, and oligodendrocytes associated with AD. A recent single-cell RNA-sequencing study found that within neurons, increased *APOE* expression leads to increased *MHC-I* expression and eventual cell death^18^. Previously, we generated snRNA-seq data from the parietal cortex from a *PSEN1* p.A79V mutation carrier and found a reduced proportion of excitatory neurons compared to two sAD samples, but a similar distribution of inhibitory neurons^19^. These studies highlight the potential for snRNA-seq to provide highly detailed expression profiles at the cellular level in a wide array of genetically defined individuals, uncovering novel cell-specific effects of AD risk genes.

In this study, we selected brains from the Dominantly Inherited Alzheimer Network (DIAN) and Charles F. And Joanne Knight Alzheimer Disease Research Center (Knight ADRC) biobanks to enrich our cohort with carriers of pathogenic, risk, and resilience genetic variants to leverage the naturally occurring perturbations of these genes and elucidate their role in AD pathogenesis. Specifically, we performed snRNA-seq utilizing the parietal cortex (Brodmann area 1-3,7) to investigate the cellular and transcriptional profiles in carriers of AD-risk-modifying variants. We focused on the parietal cortex: an understudied brain region with high level of plaques and tangle burden but relatively low atrophy in the early stages of AD pathology progression^20^. After stringent filtering, we obtained 294,114 high-quality nuclei from 67 individuals. We investigated the cellular diversity and proportions in AD by iteratively extracting nuclei by the cell type from the dataset (digital sorting), subclustering these nuclei into cell-type transcription states (cell states), and identifying differentially expressed genes (DEGs) among these cell states. We analyzed the differences in cell-type composition associated with ADAD, *APOE*ε4, *TREM2*, and *MS4A* carriers, and identified DEGs between the genetic factors and control samples. Finally, we created an online browser (http://ngi.pub/SNARE/) for convenient access to the snRNA-seq expression data.

To further investigate and validate our initial findings, we integrated brain snRNA-seq data from the dorsolateral prefrontal cortex of human donors from the ROSMAP cohort and 5xFAD mouse model (see details in Methods). We confirmed similar upregulated gene signatures between our cohort and the replication data in specific cell states of interest. Finally, we performed cell state proportional analyses to validate altered proportions associated with genetic factors.

Our findings highlight the power of leveraging genetic and single-cell molecular data to understand the heterogeneity of pathways, biological processes and cell types modulated by genetic risk factors for AD.

## Results

### Diverse transcriptional profiles within each cell type in cortical samples from genetically defined AD patients

To study the cellular and genetic heterogeneity of cortex from AD patients, we selected 74 participants from the DIAN and Knight ADRC brain banks, which included detailed clinical data, post-mortem pathological data, and genetic characterization. We generated whole transcriptomes of the parietal cortices at a single-cell resolution using 10x Genomics Next GEM technology. After data cleaning and quality control (QC; see Methods), we retained 67 samples and obtained molecular data for 16 carriers of pathogenic mutations in *APP* and *PSEN1* (autosomal dominant AD; ADAD), 19 carriers of AD risk variants in *TREM2*, 16 sAD non-carriers of any of these variants, three presymptomatic samples that meet AD pathological criteria but without cognitive impairment at age-of-death (presymptomatic), eight samples with non-AD neurodegenerative pathologies (others), and nine samples without neurodegenerative pathology nor deficits in their cognition (controls) (Table 1). Within the cohort, 41 samples carried the minor allele (A) for rs1582763, a SNP in the *MS4A* gene cluster^11,13,14^, and 24 samples carried the *APOE*ε4 allele (Table 1, Fig. 1a).

**Table 1:**
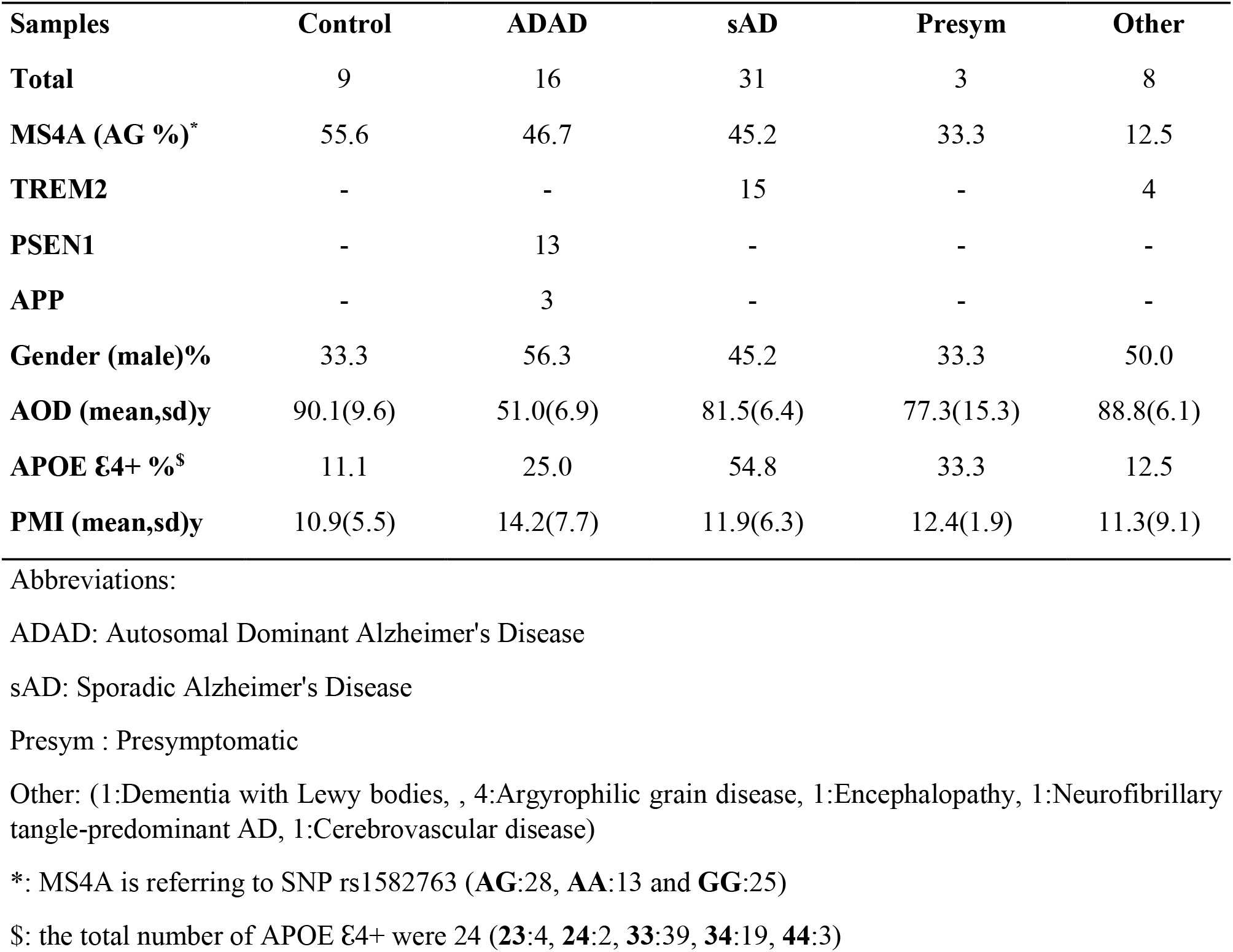
Demographic Characteristics of Samples.

**Figure 1.**
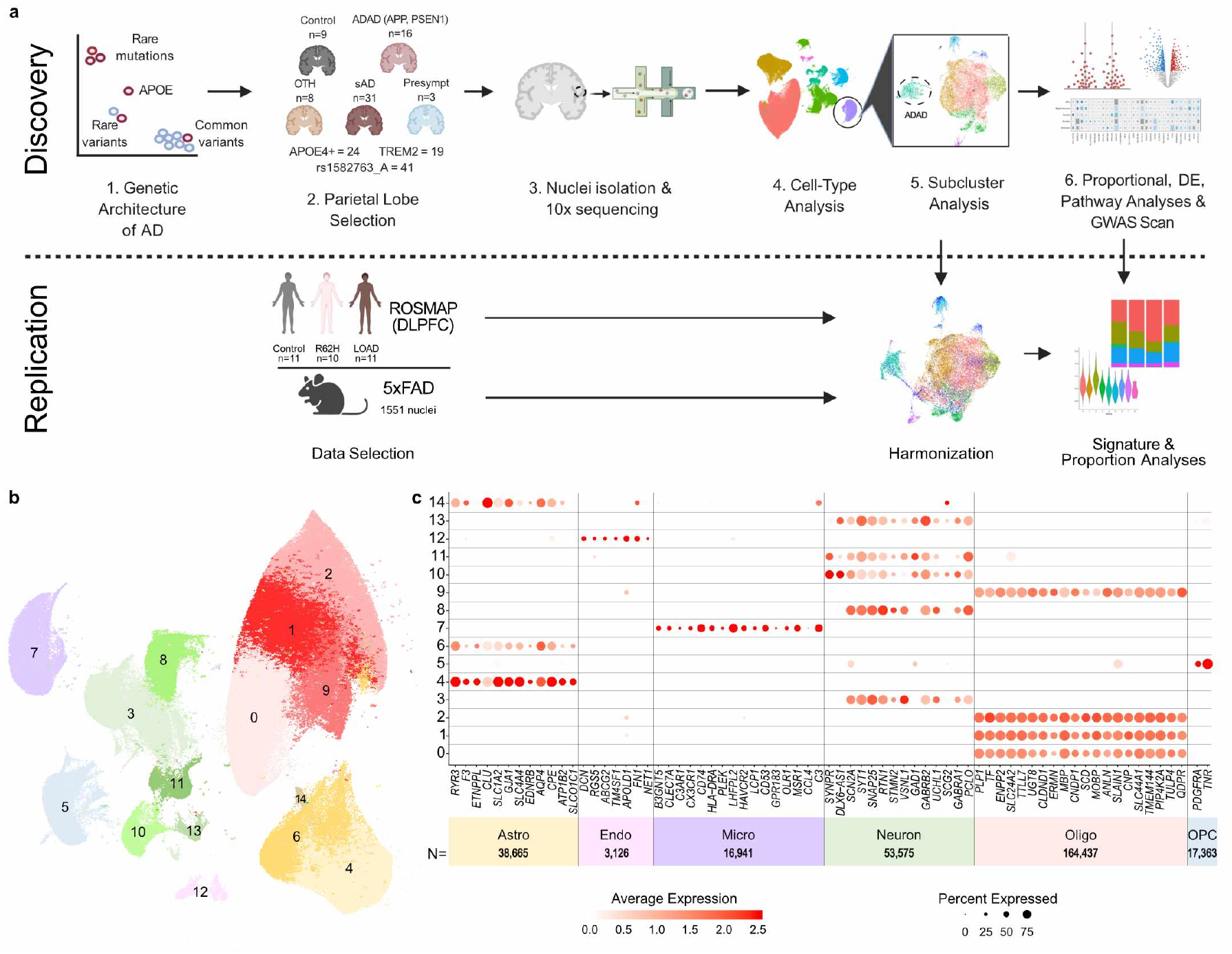
SnRNA-seq distinguishes major cell types using 67 Human brains. **a)** Diagram of the study design. **b**) UMAP plot showing 15 distinguished clusters, 0–14, with 294,114 total cells. **c**) DotPlot depicting expression of cell type specific markers genes to identify each cluster in **b**.

In total, we sequenced 1,102,459 nuclei. After data cleaning and QC (Table 2), we retained and clustered 294,114 nuclei (Table 3), identifying 15 clusters covering major cell types found in the brain (Fig. 1b,c). These clusters were annotated based on the expression of well-known cell-type markers^19^ (Fig. 1c, and Supplementary Table 1). Clusters 0, 1, 2, and 9 were identified as oligodendrocytes, clusters 3, 8, 10, 11, and 13 as neurons, clusters 4, 6, and 14 as astrocytes. Microglia, oligodendrocyte precursor cells (OPCs), and endothelial cells were identified in clusters 7, 5, and 12, respectively (Fig. 1b,c). All samples were equally distributed among the different clusters (Supplementary Table 1). The distribution of nuclei by clusters (Table 3) showed that glial cells accounted for 81.9% of the nuclei, whereas neuronal nuclei accounted for 18.1%. We observed a significant increase of OPCs (β=0.15; P=2.99×10^−2^) and endothelial cells (β=0.07; P=1.25×10^−2^) in ADAD participants compared to controls.

**Table 2:** Quality Control Steps.

|  | **Cell**<br>**ranger** | **Sigmoid**<br>**correction** | **Distrib**<br>**UMI** | **Distrib**<br>**Genes** | **% mit.** | **>10 cells\*** | **Double**<br>**Finder** |
| --- | --- | --- | --- | --- | --- | --- | --- |
| **Total #Nuclei** | 1,102,459 | 566,486 | 423,937 | 414,303 | 380,625 | 380,625 | 346,264 |
| **Mean #Genes** | 3,154 | 2,418 | 2,242 | 2,228 | 2,234 | 2,234 | 2,221 |
| **Mean #UMI** | 10,279 | 5,646 | 4,969 | 4,899 | 4,897 | 4,897 | 4,888 |
| **Avg %Mitochondria** | 2.35 | 2.34 | 1.98 | 1.94 | 1.22 | 1.22 | 1.21 |

Abbreviations:
#: number

**Table 3:** Cell Type Details.

| Cell-type | Subclusters | Nuclei | Avg DEG |
| --- | --- | --- | --- |
| Oligodendrocytes | 9 | 164,437 | 3,329 |
| Neurons (Ex/Inh) | 4/3 | 38,164/15,411 | 4,713/3,881 |
| Astrocytes | 5 | 38,665 | 3,326 |
| OPC | 7 | 17,363 | 1,962 |
| Microglia | 9 | 16,941 | 1,172 |
| Endothelial | -- | 3,126 | -- |
| **Total** |  | **294,114** |  |

Ex: Excitatory Inh: Inhibitory

After extracting each cell type and subclustering, we identified 5-9 subclusters or cell-type transcription states (cell states), within each cell type (Table 3). We observed an average of 2,696 upregulated genes (average of 16,223 tested) that passed multiple test correction in each cell state compared to all other cell states in the same cell type (Supplementary Table 3, Supplementary Material). This highlights the incredible multiplicity within cell types. As expected due to statistical power, there was a positive correlation between the number of detectable DEGs and the number of nuclei in the cell type. However, the highest average number of DEGs was found in inhibitory and excitatory neurons, which had three times less nuclei than that of oligodendrocytes, most likely due to the neuronal differences between cortical layers^21^. We identified a comparatively small number of endothelial nuclei, which were not further subclustered (Table 3).

### Pleiotropic effects of *APP* and *PSEN1* mutations across brain cell-types

The core of the amyloid hypothesis is that ADAD mutations in *APP* and *PSEN1* lead to dysfunctional neurons, the main source of aggregation-prone Aβ. Strikingly, every cell type we analyzed exhibited cell states enriched within ADAD carriers (Fig. 2a-e and Supplementary Table 4). Two astrocyte cell states were specific to ADAD subjects. One in particular, Astro.4 (β=0.15, P=4.39×10^−2^), had increased expression of *OSMR* (log2FC=1.48, Adj.P=4.37×10^−4^), *VIM* (log2FC=1.80, Adj.P=8.92×10^−13^), and *CTSB* (log2FC=1.56, Adj.P=4.03×10^−5^) compared to the other astrocyte cell states, similar to the disease-associated astrocytes (DAA) identified in the 5xFAD mouse model^22^ (Extended Fig. 1a).

**Figure 2.**
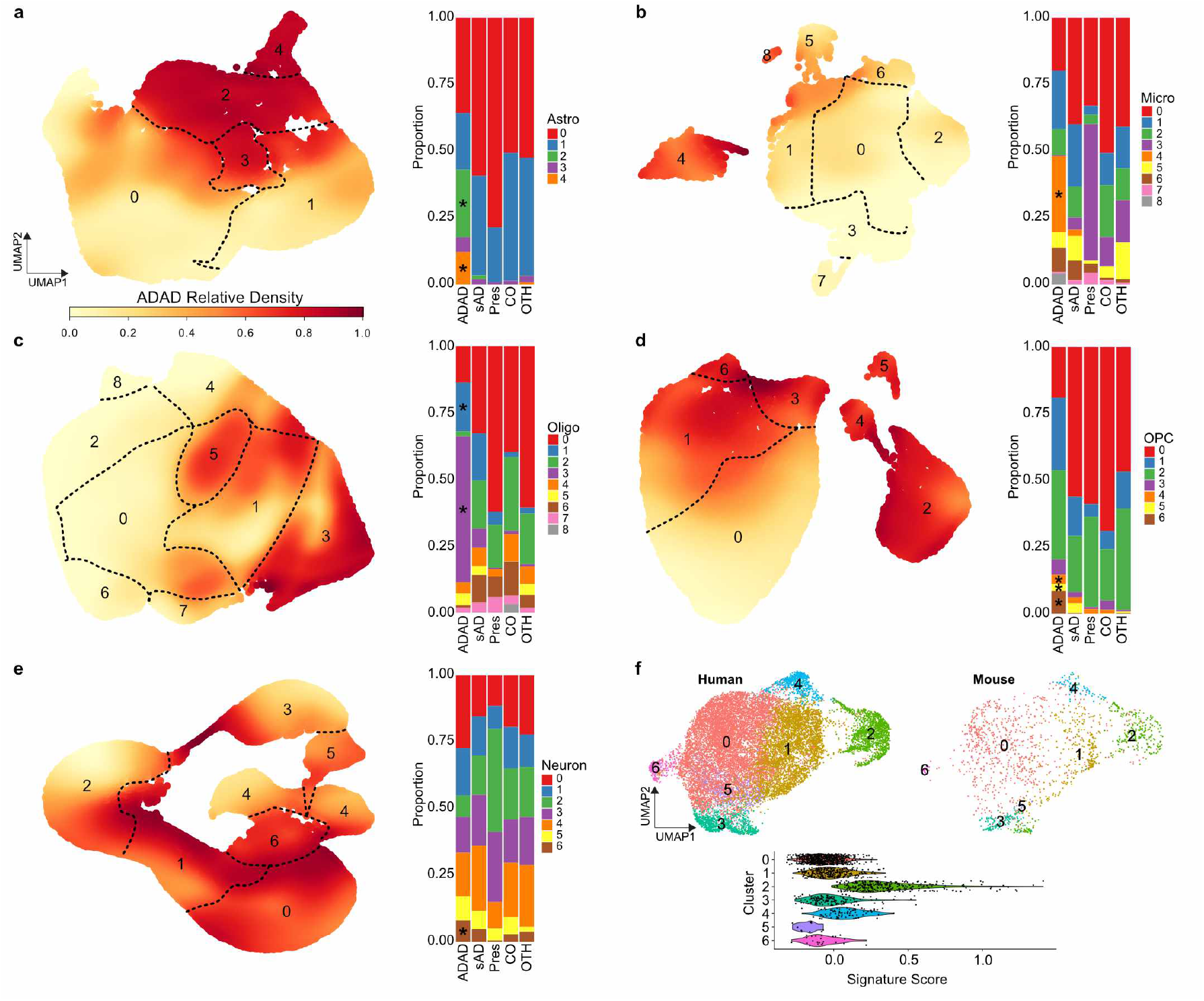
ADAD participants exhibit distinct signatures in Astrocytes, Microglia, OPCs, Oligodendrocytes, and Neurons. **a - e)** Left panel, UMAP Enrichment plots show the normalized density of ADAD nuclei by cell type. 0 and 1 represent the lowest and highest measured normalized densities respectively. Right panel, a proportion plot shows the enrichment of certain cell states in ADAD participants. (*) represent a significant (P<0.05) enrichment. ADAD = autosomal dominant AD; sAD = sporadic AD; Pres = presymptomatic; CO = neuropath free; OTH = non-AD neurodegenerative. **a)** Astrocytes. **b)** Microglia. **c)** Oligodendrocytes. **d)** OPCs. **e)** Neurons. **f)** 5xFAD mouse validation of microglia ADAD cluster (cluster 4 in **b**, cluster 2 here). On the top, a UMAP of integrated microglia split by species. On the bottom, a violin plot showing that mouse cells in the ADAD cluster have a higher human microglia ADAD cluster signature score than cells in other clusters.

Microglia contained a prominent cell state, Mic.4 (β=0.40, P=2.50×10^−3^), composed of nuclei from 11 *PSEN1* carriers and one *TREM2* p.R136W carrier (Fig. 2b). This cell state did not show a significant overlap of DEGs with genes featured in established microglia signatures including ‘disease-associated microglia’ (DAM)^23^, ‘microglial neurodegenerative’ (MGnD)^24^, and ‘human AD microglia’ (HAM)^25^ (Supplementary Table 5). Interestingly, Mic.4 showed a significant increase in the expression of *MECP2* (β=0.67, Adj.P=3.20×10^−10^), which has previously shown differential expression in AD brains and when knocked down in microglia, caused NMDA receptor-dependent excitotoxic neuronal cell death in a mouse model of Rett syndrome^26^. Pathway analysis revealed that the 491 upregulated genes in Mic.4 were associated with MAPK signaling (Adj.P=1.37×10^−2^), estrogen signaling (Adj.P=1.37×10^−2^), NOD-like receptor signaling (Adj.P=1.90×10^−2^), and necroptosis pathways (Adj.P=2.42×10^−2^), indicating an acute cellular response leading to cell death (Supplemental Table 6). Furthermore, we also observed this microglia cell state in the cortex of seven-month-old 5xFAD mice (high amyloid plaque load at this age), with the same upregulated gene signature (upregulation signature score P=1.92×10^−374^; Fig. 2f).

Oligo.3 was also significantly associated with ADAD subjects (β=0.63, P=1.48×10^−6^). Pathway analysis of the oligo.3 upregulated genes revealed an enrichment in spliceosome genes (Adj.P=4.64×10^−24^) many of which were from the family of heterogeneous nuclear ribonucleoproteins (HNRNP), including *HNRNPA1, HNRNPA2B1, HNRNPA3, HNRNPC, HNRNPD, HNRNPH3, HNRNPK, HNRNPM*, and *HNRNPU*, which have previously been linked to late-onset AD^27^ (Extended Fig. 1b, Supplementary Table 6h). The AD risk genes, *PICALM, CLU, APP*, and *MAP1B*, that have intronic excision levels correlated with the expression of HNRNP genes^27^, were also overexpressed in Oligo.3 (Extended Fig. 1b). This suggests that *PICALM, CLU, APP*, and *MAP1B* could be alternatively spliced through HNRNP splicing repression within oligodendrocytes. HNRNP genes also play a role in amyotrophic lateral sclerosis (ALS) and frontotemporal dementia (FTD)^28^ and promote translation of APP^29^. Similarly, additional clusters in oligodendrocytes (Oligo.1), OPCs (clusters 4, 5, 6), and Neurons (Neuro.6) were enriched within ADAD carriers (Fig. 2c-e).

### *TREM2* variants modulate microglial and oligodendrocytic transcription states

Individuals with specific *TREM2* AD risk variants (p.R47H, p.R62H and p.H157Y) showed increased proportions of nuclei in Mic.2 (Fig. 3a; β=0.23, P=3.29×10^−2^). Mic.2 showed high expression of resting-state-microglia marker genes (*TMEM119, P2RY13, MED12L* and *SELPLG*) and minimal elevation of activated (*ABCA1, C5AR1, TNFAIP3*, and *CD83*) marker genes compared to our resting (Mic.0) and activated (Mic.1) microglia cell states (Fig. 3b). This is consistent with the reported association of these *TREM2* risk variants with reduced microglial activation^16,30^. To replicate the enrichment of this cluster within *TREM2* risk variant carriers, we analyzed microglia from 32 ROSMAP snRNA-seq samples (DLPFC) that included 11 *TREM2* p.R62H carriers (Synapse ID: syn21125841)^16^. We found that 10.2% of the ROSMAP microglia recapitulated the Mic.2 transcriptional state (signature score: β=0.22 P=7.4×10^−107^), and that on average, carriers of *TREM2* p.R62H had a higher proportion of their microglia in this unique state than non-carriers (Fig. 3a, Discovery: β=0.20, P=2.58×10^−2^; Meta-analysis: P=2.26×10^−2^).

**Figure 3.**
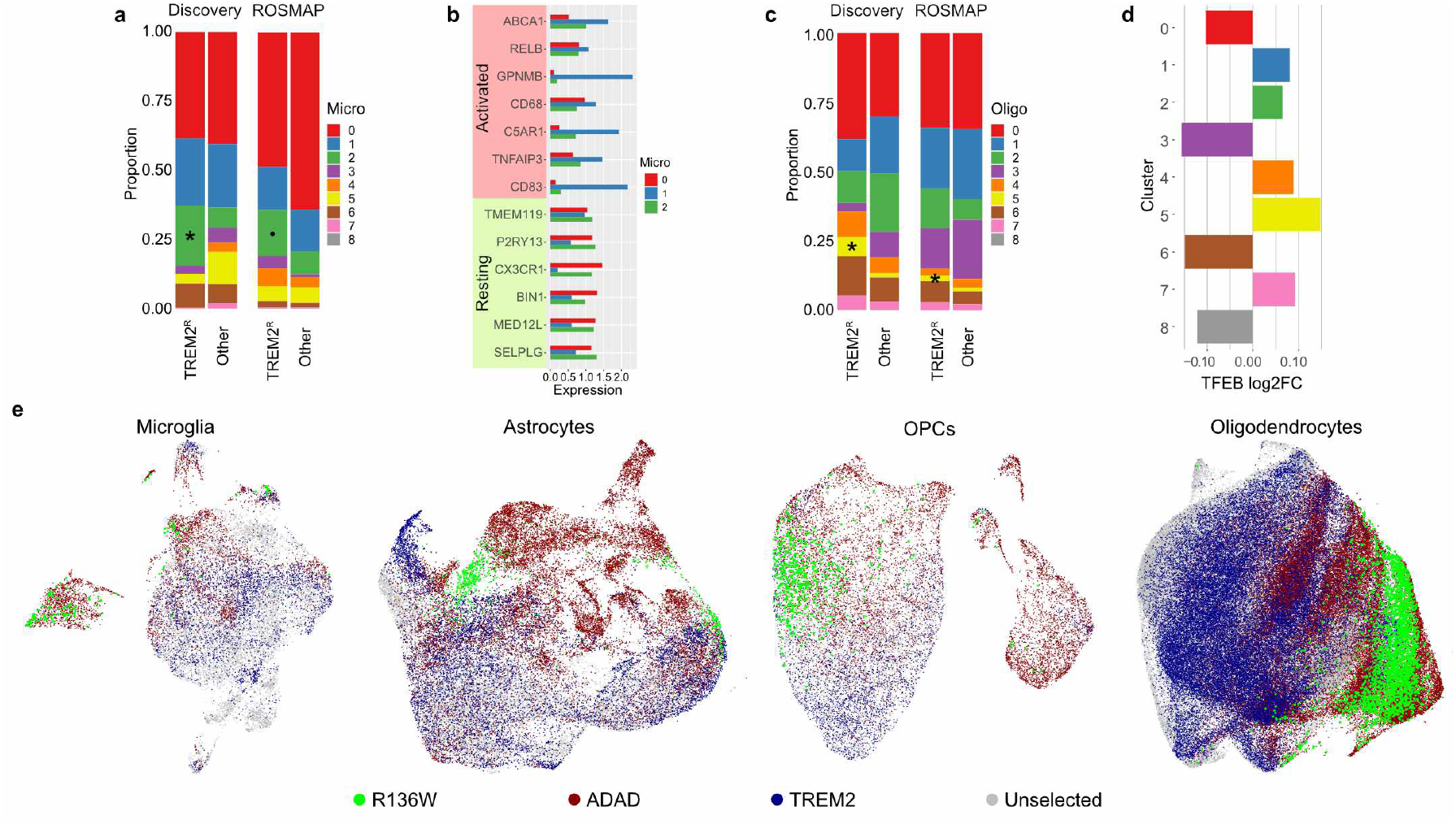
*TREM2* reduced-activation variant carriers (p.R47H, p.R62H, and p.H157Y) have distinct microglia and oligodendrocyte profiles. **a)** Proportion plot shows a significant (P<0.05) enrichment (*) of mic.2 in *TREM2* reduced activation carriers (TREM2^R^) and a trend (**·**) in the ROSMAP replication. ‘Other’ indicates all sAD non-*TREM2* reduced activation carriers including carriers of other *TREM2* variants. **b)** Bar plot shows the expression of both activated and resting microglia marker genes in mic.2 compared to the resting (mic.0) and activated cell states (mic.1). **c)** Proportion plot shows a significant (P<0.05) enrichment (*) of oligo.5 in TREM2^R^ carriers in both the discovery and ROSMAP cohorts (P<0.05). **d)** Bar plot shows the log2 fold change of *TFEB* by oligodendrocyte subcluster. **e)** *TREM2* p.R136W carrier (green) clusters more closely with ADAD (red) than other *TREM2* carriers (blue) in Microglia, Astrocytes, OPCs, and Oligodendrocytes (from left to right).

These *TREM2* risk variant carriers were also enriched for Oligo.5 (β=0.13, P=4.66×10^−2^; Fig. 3c; Supplementary Table 4b), which exhibited upregulation of 1,124 genes including *TFEB* (Log2FC=0.15; Adj.P=8.69×10^−6^) compared to other oligodendrocyte cell states (Fig. 3d). Altered *TFEB* expression may be driven by the interaction of *TREM2* with *mTOR* which is upstream of *TFEB*^*31,32*^. *TFEB* is a central regulator of lysosomal biogenesis^33,34^ and represses myelination at different developmental stages^35^. Altered *TFEB* signaling has been implicated in a number of neurodegenerative diseases^33,36^. Oligo.5 also had increased proportions among sAD subjects (Supplementary Table 4d). This cell state was also present in 7.1% of oligodendrocytes from the ROSMAP cohort (upregulation signature score of Oligo.5 p=9.71×10^−96^) with an increased proportion in p.R62H carriers (Discovery: β=0.13, P=2.48×10^−2^; Meta-analysis: P=6.11×10^−3^) (Fig 3c). Changes in autophagy markers were reported previously in microglia of p.R62H carriers^31^ and our data suggest that autophagy dysregulation may also be a feature of oligodendrocytes in p.R62H carriers.

An inspection of the *TREM2* p.R136W variant^10^ carrier revealed a divergent signature compared to other *TREM2* carriers. Cellular studies showed that *TREM2* p.R136W severely altered both cell surface and overall TREM2 expression^37^. Three *TREM2* variant carriers, including p.R136W, were diagnosed with AD prior to age 65, but only the p.R136W carrier had a Braak stage of VI (6). In each cell type, this individual’s nuclei tended to cluster with the ADAD nuclei (Fig 3e), suggesting that this mutation could altered expression networks more similarly to *PSEN1* or *APP* mutations than to other *TREM2* variants. Due to the rarity of this variant, only a single brain was analyzed; thus, additional replication is required to validate these observations.

### MS4A resilience variant carriers show a specific inflammatory microglial activation state

Carriers of rs1582763-A, an intergenic allele associated with reduced risk for AD and higher CSF sTREM2 levels^14^, exhibited a trend for decreased proportions of nuclei in Mic.1 (Supplementary Table 4c), an activated microglia state. Mic.1 had an upregulation of activated-microglia genes *CD68* (log2FC=0.39, P=1.06×10^−4^), *CD83* (log2FC=1.34, P=9.44×10^−37^), *TNFAIP3* (log2FC=0.57, P=2.16×10^−7^), *C5AR1* (log2FC=1.23, P=4.03×10^−46^), *GPNMB* (log2FC=1.73, P=6.99×10^−102^), and *ABCA1* (log2FC=0.82, P=1.78×10^−45^; Fig. 3b), and a significant overlap with genes also upregulated in the DAM^23^, MGnD^24^, ‘human Alzheimer’s microglia’ (HAM)^25^, and aging^38^ signatures (Supplementary Table 5). In contrast, rs1582763-A carriers showed increased proportions of nuclei in Mic.3 compared to non-carriers (β=0.15, P=1.67×10^−3^; Fig 4a,b; Supplementary Table 4c). The Mic.3 state displayed a proinflammatory profile, including the upregulation of *ILB1* (log2FC=3.45, Adj.P=2.07×10^−36^), *CD14* (log2FC=0.63, P=1.87×10^−3^), *FCGR3A* (log2FC=0.50, P=3.31×10^−3^), and *CD40* (log2FC=0.91, P=9.84×10^−3^; Fig. 4c). Although this rs1582763-associated microglia cell state was present in the ROSMAP cohort (upregulation signature score of Mic.3 P=8.64×10^−152^), it was not significantly associated with rs1582763, possibly because the ROSMAP cohort contained few homozygous recessive carriers (n=3) which were enriched by the design of our study.

We then performed differential expression analyses to characterize the genes that differentiated the main activated microglia (Mic.1) from the rs1582763-A activated microglia (Mic.3). Mic.1 showed increased expression of *C5* (log2FC=1.79, Adj.P=3.46×10^−3^), whereas Mic.3 had increased expression of *C3* (log2FC =-1.24, Adj.P=4.98×10^−26^; Fig. 4d; Supplementary Table 7a). Previous analyses have indicated a protective role for *C3* in AD and a potentially detrimental role for *C5*^39^. This is consistent with the protective effect of rs1582763-A. Mic.1 also had increased *ACVR1* (log2FC =0.97, Adj.P=2.92×10^−3^) and *BMPR2* (log2FC =0.39, Adj.P=3.54×10^−2^),indicating increased BMP signaling, whereas Mic.3 had high *TGFBR1* (log2FC =-0.98, Adj.P=4.82×10^−17^) and *TGFBR2* (log2FC =-0.35, Adj.P=4.50×10^−2^), indicating increased TGF-β signaling (Fig. 4d, Supplementary Table 7b). These genes encode for related receptors in the TGF-β superfamily which is implicated in multiple neurological disorders^40^. A gene ontology analysis also showed an upregulation of genes related to cytokine response/production (Adj.P=2.89×10^−9^) and response to lipopolysaccharides (Adj.P=1.78×10^−4^). In addition, carriers of rs1582763-A had decreased proportions of resting astrocytes (Astro.0) and a trend towards increased proportions of activated astrocytes (Astro.1) (Fig. 4e, Extended Fig. 1a, Supplementary Table 4). The MS4A genes are not expressed in astrocytes which suggests a possible cellular cross-talk synchronizing the activation of microglia and astrocytes.

**Figure 4.**
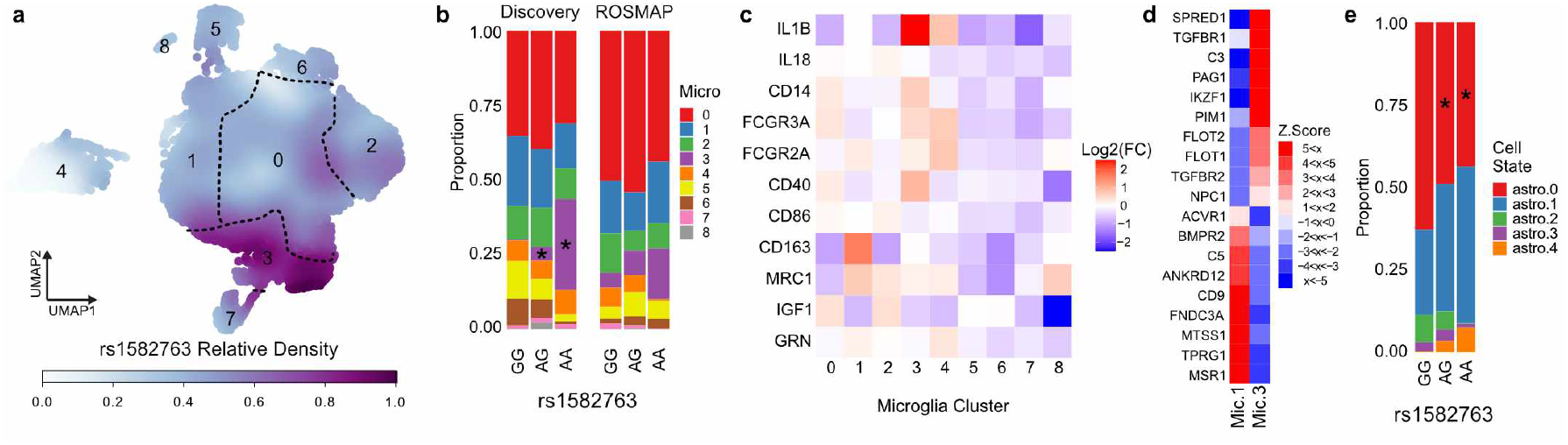
Unique microglia signatures for *MS4A* rs1582763 carriers. **a)** UMAP Enrichment plots show the normalized density of rs1582763_A alleles in microglia. 0 and 1 represent the lowest and highest measured normalized densities respectively. **b)** A proportion plot shows a dose dependent enrichment of microglia expression state 3 in rs1582763 carriers. (*) represents a significant (P<0.05) enrichment and (**·**) represents a trend. **c)** A heatmap of proinflammatory (*IL1B* – *CD86*) and anti-inflammatory (*CD163* – *GRN*) microglia marker gene log2 fold change (Log2(FC)) within the microglia subclusters. **d)** A heatmap of linear regression z scores shows the differences in gene expression between the ‘main’ (Mic.1) and ‘MS4A’ (Mic.3) activated microglia clusters. **e)** A proportion plot depicts a significant (P<0.05) depletion (*) of astro.0 (non-activated) in rs1582763 carriers and a trend for enrichment of astro.1 (activated).

### Coexpression of *APOE* and *MHC-I* highlights inhibitory neuron vulnerability to neurodegeneration

*APOE*ε4 carriers did not exhibit a unique cell state signature within any cell type likely because we did not enrich the data set for this allele (Table 1, Supplementary Table 4e). However, *APOE*ε4 carriers showed an upregulation of genes involved in ribosome biogenesis (Adj.P=1.42×10^−2^) and mitotic G1/S transition checkpoint signaling (Adj.P=1.42×10^−2^) in microglia, maintenance of protein localization in ER (Adj.P=7.75×10^−3^) and COPI coating of Golgi vesicles (Adj.P=7.75×10^−3^) in astrocytes, and cytoplasmic translation in both neurons (Adj.P=3.24×10^−9^) and OPCs (Adj.P=4.34×10^−5^; Supplementary Table 8). We also explored the role of *APOE* expression in neurons. As was previously described^18^, we found a moderate proportion of *APOE*-high expression neurons in controls (20%) with significantly higher proportions in presymptomatic brains (69%, P=6.52×10^−4^) and considerably lower proportions in both sAD (5.7%, P=2.91×10^−2^) and ADAD (3.4%, P=2.20×10^−2^) brains (Fig. 5a,b). We also observed a correlation between *APOE* and *MHC-I* within specific populations of neurons (Fig. 5c-h). Only inhibitory neurons (Neuron.3 and Neuron.5) showed significant correlations in almost all sub-groups (ADAD, sAD, presymptomatic, and controls), but all inhibitory and excitatory cell states had substantial correlations in ADAD participants (Fig. 5d-h, Supplementary Table 9). An increased expression of *APOE* and *MHC-I* within a neuron seems to tag it for removal^18^, which agrees with the sharp increase in *APOE-*high neurons prior to clinical manifestations (presymptomatic) of AD followed by a sharp decrease after (sAD and ADAD). The correlation in Neuron.3 and Neuron.5 across sub-groups supports the premise that these inhibitory neurons are the first to succumb to neurodegeneration^41^ and that as pathology progresses, excitatory neurons are also targeted, as seen in the ADAD participants. *APOE*-high nuclei in Neuron.3 and Neuron.5 had upregulated genes in ‘cellular response to cytokine stimulus’ (Adj.P=3.24×10^−10^), suggesting cytokines could trigger the increased *APOE* and *MHC-I* expression in these cells^42^ (Supplementary Table 10a). Dysregulated genes were involved in circadian entrainment, cAMP signaling, and NMDA receptor activity as previously reported and highlight sleep and circadian rhythm disruption in AD^43^ (Supplementary Table 10b).

**Figure 5.**
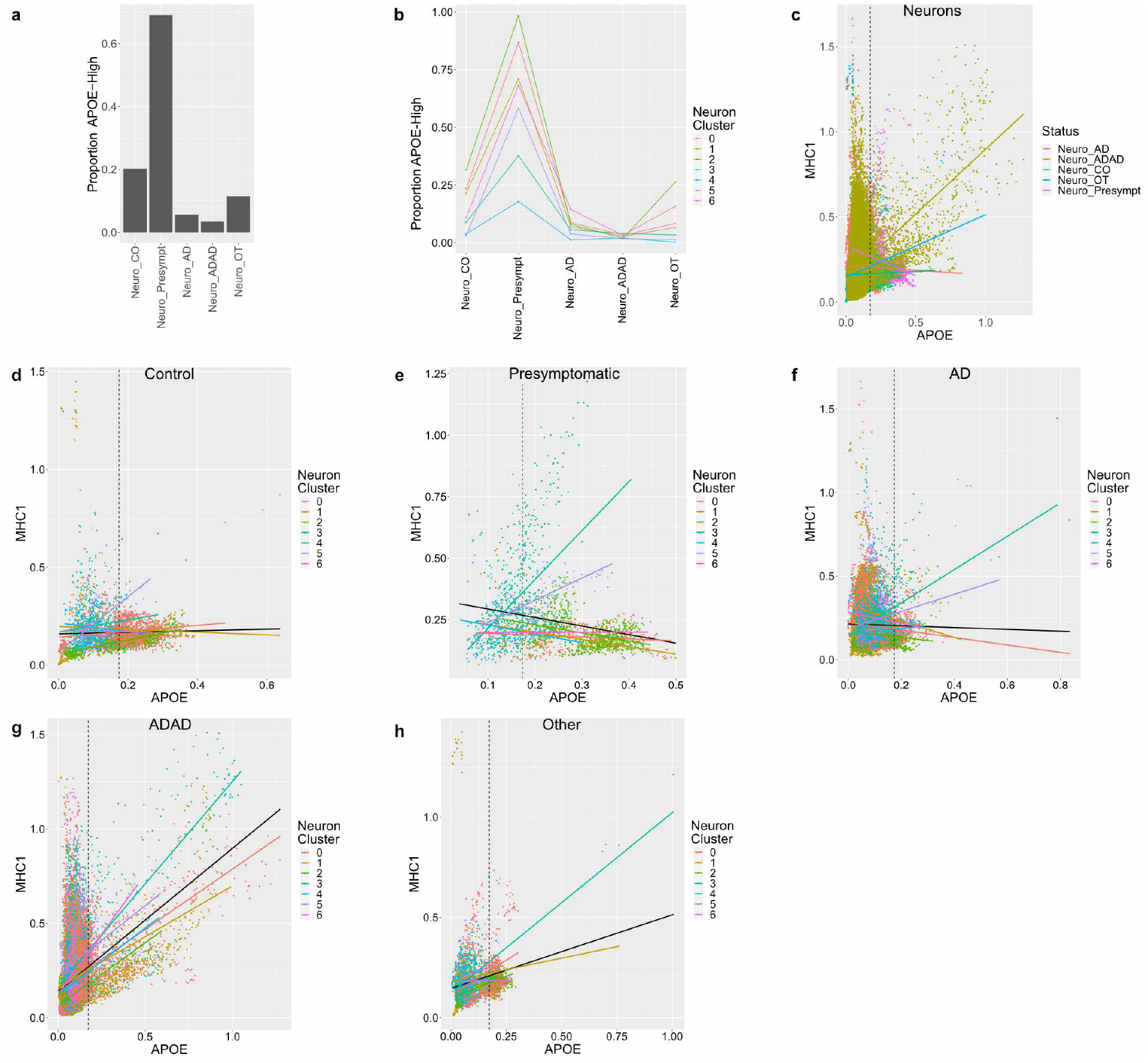
Neuronal APOE-high expression patterns. **a)** Bar plot showing the average proportion of APOE-high neurons in each status. **b)** Average proportion of *APOE*-high neurons in each status split by subcluster. Clusters 0, 1, 2, 6 are excitatory and clusters 3, 4, 5 are inhibitory. **c-h)** Scatter plots and regression lines depicting the coexpression between *APOE* and *MHC-I* in neurons after MAGIC imputation. The vertical dotted line marks the *APOE*-high threshold. **d-h)** The black regression line depicts the overall correlation within the specified status.

### Genes within risk AD GWAS loci show variable expression between cell states

GWAS meta-analyses have successfully identified genomic loci associated with sAD^11-13^. However, several major hurdles have prevented the field from understanding the contribution of these genomic loci to AD risk. For example, the cell types in which the genomic loci are expressed and their function/dysfunction in these cell types is poorly understood. We sought to leverage our glial and neuronal cell states to map genomic loci implicated in the general AD population onto genes and cell types. We curated a list of genes in loci identified in recent AD GWAS, starting with 89 genes previously prioritized^*13,44,45*^ (77 measured in this study) from 39 different loci (Supplementary Table 11a); we extended the list by adding the ‘non-prioritized’ genes within all AD GWAS associated loci also present in this data set (540 genes, 46 loci; Supplementary Table 11b,c).

We then inspected the snRNA-seq data to identify which genes were differentially expressed among cell states. Out of the 77 prioritized genes, 39 (from 29 loci) showed differential expression between the cell states of at least one cell type (Fig. 6). We also found that for 13 genes, the cell type with the greatest expression level variability (log fold change between cell states) did not match the cell type with the highest average expression. For example, *BIN1* showed the highest average expression in oligodendrocytes, but OPCs had the largest log2 fold change in expression (log2FC) between cell states. This suggests that transcriptional states can provide additional independent information to map GWAS genes to cell types. Rare coding variants in *PLCG2*, implicated in *TREM2*-dependent microglial function in AD^46^, have been recently reported to protect against AD^47^. We found differential expression of *PLCG2* between cell states within all cell types, suggesting an extended functionality beyond that of the *TREM2* signaling pathway in microglia (Fig 6)^48,49^. *PLCG2* expression was significantly increased in ADAD brains across all cell types except microglia, which was only nominally significant (Supplementary Table 12). *SORL1*, predominantly studied in neurons in the context of AD^50^, showed higher average expression in microglia than in neurons and greater log2FC within oligodendrocytes than in neurons. Finally, we report that the three prioritized genes in the *EPHA1* locus (*EPHA1, ZYX*, and *FAM131B*) showed low average expression and minimal log2FC in all cell types (Fig 6). However, when we included all genes in the locus, we found moderate average expression and significant log2FC of *TCAF1* in all cell types except microglia suggesting that this gene could be the lead gene at this locus (Supplementary Material). *TCAF1* helps traffic the receptor *TRPM8* to the plasma membrane, where it regulates inflammation^51,52^. An alternative explanation is that the effector cell type was not well represented in the present study (e.g. endothelial cells, pericytes). These results will be valuable in identifying the relevant genes and cell types linked to GWAS loci.

**Figure 6.**
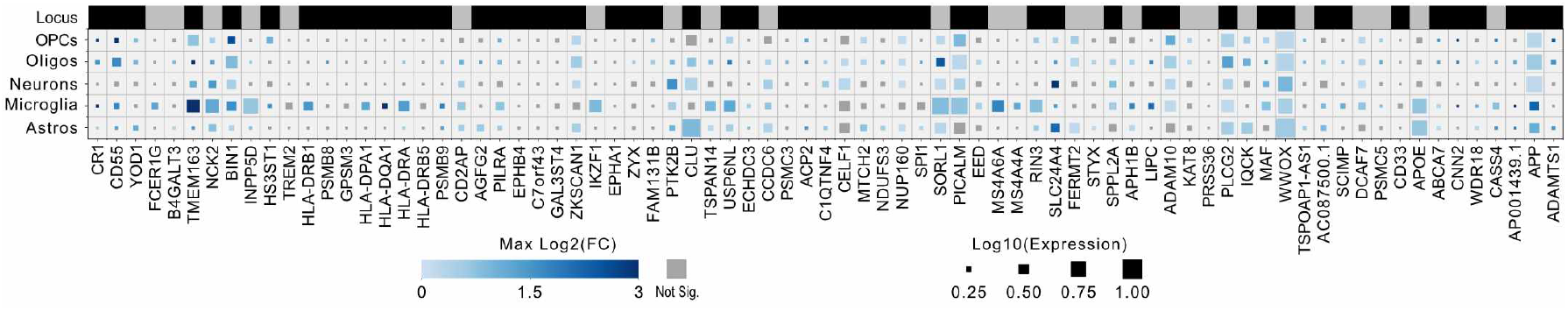
Cell type specific expression and cell state differential expression of prioritized genes help link GWAS hits to cell types. A modified heatmap shows the average expression and max log2 fold change within cell types of prioritized genes in GWAS loci. At the top, the ‘Locus’ row indicates genes within the same locus using alternating black and gray rectangles. In the body of the plot, the color of the squares represents the max log2 fold change of the gene between cell states (subclusters) of that cell type. The log2 fold changes greater than three were set to three to preserve visual variability. Gray squares indicate no significant log2 fold change within that cell type. Square size represents log10 transformed gene expression. 77 of the 89 genes prioritized by Kunkle *et al*, Andrews *et al*. and Schwartzentruber *et al*. passed QC in our data.

## Discussion

We performed snRNA-seq on over one million nuclei from the parietal cortex (area B1-3,7) of 74 brains with AD genetic variants in *APP, PSEN1, TREM2*, and rs1582763-A, a resilience intergenic variant in the *MS4A* locus associated with increased levels of soluble TREM2 in CSF and reduced AD risk^14^. After a stringent QC process, we retained 67 brains and 294,114 high-quality nuclei representing most major cortical cell types. Our high numbers of nuclei enabled re-clustering of the individual cell types to identify unique biological cell states.

One of the premises of the amyloid hypothesis is that the molecular changes in ADAD could be extrapolated to sAD. Interestingly, we observed cell states that were either unique to or enriched within ADAD subjects for all cell types. Thus, one could conclude that ADAD has altered cellular architecture compared with sAD invalidating extrapolation. However, upon integration of our microglia and oligodendrocyte data from the parietal cortex with snRNA-seq data from the DLPFC of ROSMAP participants, we observed ROSMAP (sAD) nuclei clustering with our ADAD enriched clusters despite the absence of ADAD subjects in the ROSMAP cohort (Extended Fig. 2b,e). The parietal cortex is affected in the later stages of AD progression, whereas the DLPFC is affected earlier in disease progression^53,54^. Therefore, the cell states identified in the parietal cortex of ADAD participants could represent advanced/accelerated stages of pathology which can also be detected in brain regions with high pathology in LOAD like the DLPFC. This would be consistent with the elevated tau PET found in the parietal region of ADAD compared to sAD patients^55^. The differences observed in the parietal cortex could also be due to younger ages in ADAD (average of 32.9 years younger). Additional studies capturing multiple brain regions with varying degrees of pathology and ages are required to understand these observations in more detail.

We report microglia and oligodendrocyte cell states associated with *TREM2* risk variant carriers. *TREM2* is mainly expressed in microglia. However, loss of function mutations in TREM2 cause Nasu-Hakola disease, which is characterized by white matter changes, including loss of myelination, suggesting an oligodendrocyte-microglia crosstalk^56^. Therefore, it is likely that AD patients with *TREM2* variants had altered microglia behavior which changed the microenvironment, driving the oligodendrocyte cell state found in *TREM2* variant carriers. The microglial cluster associated with these reduced activation variant carriers displayed a resting-like state, suggesting they might benefit from antibody treatments targeting *TREM2* to prevent its cleavage, increasing microglial activation^57^. However, not all *TREM2* risk variants are functionally equivalent^14,58,59^. *TREM2* p.R47H, p.R62H, and p.H157Y show reduced cell activation *in vitro*^16,30^ and we demonstrate that brains from these variant carriers exhibit altered microglia and oligodendrocytes populations. Interestingly, a single *TREM2* p.R136W carrier clustered closer with the ADAD subjects than the other *TREM2* carriers in astrocytes, microglia, oligodendrocytes, and OPCs.

We also observed a microglia state expressing activation markers enriched for homozygous carriers of the *MS4A* resilient variant. In contrast to the major activated microglia state, this cell state showed upregulation of proinflammatory genes and other genes related to cytokine signaling. We also replicated the finding that the number of *APOE*-high neurons increases before the clinical onset of AD and decreases during disease progression^18^. We also found a correlation between *APOE* expression and *MHC-I* expression. This correlation was predominantly seen in inhibitory neurons, which supports the current view that inhibitory neurons are affected before their excitatory counterparts^41^. ADAD participants showed significant coexpression of *APOE/MHC-I* in all neuronal cell states.

The majority of the AD GWAS hits are non-coding variants and potentially cause altered gene expression. Thus, we inspected our cell state differential expression results to identify genes within AD GWAS loci that showed variable expression within cell types. The results highlight the specific cell types influenced by the genes and show that many genes play essential roles across cell types, including *PLCG2* and *SORL1*. In the future, cell-type-specific eQTL analyses using the lead GWAS hits could confirm if any of the expression variability is due to genotypic status.

In conclusion, a single AD risk variant can alter the transcriptional landscape of multiple brain cell types. Pathogenic mutations in *APP* and *PSEN1* influenced profiles of neurons but more especially glia when compared to controls and sAD parietal cortices. We found *TREM2* risk variants that shifted microglial and oligodendrocytic profiles and that the resilience *MS4A* variant altered the ‘activated’ microglia profile. Each of these changes has the potential to modify the pathological progression and the clinical manifestations of AD. Overall, this suggests the critical need to incorporate genetics when screening participants for drug trials and selecting the optimal treatment program for AD patients.

## Materials and Methods

### Processing of brain tissue samples

The Neuropathology Core of the Knight-Alzheimer Disease Research Center (Knight-ADRC) and the Dominantly Inherited Alzheimer Network (DIAN) provided the parietal lobe tissue of postmortem brains for each sample. These samples were obtained with informed consent for research use and were approved by the review board of Washington University in St. Louis. AD neuropathological changes were assessed according to the National Institute on Aging-Alzheimer’s Association (NIA-AA) criteria. Their demographic, clinical severity and neuropathological information are presented in Table 1.

### Nuclei isolation and snRNA-seq on the 10X Genomics platform

The 74 frozen human parietal cortices were processed according to the ‘Nuclei extraction and library preparation’ protocol described in ^19^. Briefly, the tissue was homogenized and the nuclei were isolated using a density gradient. The nuclei were then sequenced using the 10X Chromium single cell Reagent Kit v3, with 10,000 cells per sample and 50,000 reads per cell for each of the 74 samples.

### SnRNA-seq data processing with 10X Genomics CellRanger software and data filtering

The CellRanger (v3.0.2 10XGenomics) software was employed to align the sequences and quantify gene expression. We used the GRCH38 (3.0.0) reference to prepare a pre-mRNA reference according to the steps detailed by 10X Genomics. The software was packaged into a Docker container (https://hub.docker.com/r/ngicenter/cellranger3.0.2), allowing us to launch it within the McDonnell Genome Institute (MGI) infrastructure, reducing the computing time for generating the BAM files.

Filtering and QC were done using the Seurat package (3.0.1) on each subject individually. Each raw gene expression matrix for each sample was plotted, using *BarcodeInflectionsPlot* to calculate the inflection points derived from the barcode-rank distribution. Once the thresholds were determined, a subset of the data was isolated. We removed nuclei with high mitochondria gene expression following the dynamic model proposed by Tsai et al.^15^. Briefly, the nuclei were grouped by their percentage of mitochondria values using k=2 clustering, and the group with the higher percentage values was removed. Genes that were not expressed in at least ten nuclei were removed from the final matrix. To detect and discard doublets, we used *Doublet Finder*^60^, which removes nuclei with expression profiles that resemble synthetically mixed nuclei from the dataset. The gene expression matrices from all samples were combined in R independently for further processing using the Seurat protocol.

### Dimensionality reduction, clustering, and UMAP

The merged expression matrix was normalized using the *SCTransform* protocol by Seurat. This function calculates a model of technical noise in scRNA-seq data using ‘regularized negative binomial regression’ as described previously^61^. We regressed out, during the normalization, the number of genes, the number of UMIs, and the percentage of mitochondria. The principal components were calculated using the first 3000 variable genes, and the Uniform Manifold Approximation and Projection (UMAP) analysis was performed with the top 14 PCs. The clustering was done using a resolution of 0.2.

### Cluster annotation and quantification of regional and individual contributions to cell types

We employed a list of marker genes that we had previously curated^19^ to annotate brain single-nuclei RNA-seq data. We used the *DotPlot* function (Seurat package) to visualize the average expression of genes related to specific cell types. This approach enabled the labeling of cell types based on the overall expression profile of the nuclei, regardless of dropout events. In addition, we employ a supervised method termed Garnett^62^ that leverages machine learning to classify each of the nuclei and estimate cluster homogeneity. This method also provides a metric of gene ambiguity, which enables further optimization of the marker genes to be included in the classification process. For this method, we employed *SYT1, SNAP25, GRIN1* to classify neurons, *NRGN, SLC17A7, CAMK2A* for excitatory neurons and *GAD1, GAD2* for inhibitory neurons; *AQP4, GFAP* for astrocytes; *CSF1R, CD74, C3* for microglia; *MOBP, PLP1, MBP* for oligodendrocytes; *PDGFRA, CSPG4, VCAN* for oligodendrocyte precursor cells (OPCs); *CLDN5, TM4SF1, CDH5* for endothelial cells and ANPEP for pericytes. We employed the function *check_markers* (Garnett package) to evaluate the ambiguity score and the relative number of cells for each cell type. A classifier was then trained using the marker file, with “num_unknown” set to 50. This classifier annotates cells with cell-type assignments extended to nearby cells using the “clustering-extended type” labeling option. At this stage, 1 ambiguous cluster and 1 subject-specific cluster were dropped.

### Identification of alternative cell-type transcription states

The nuclei within the primary cell-type clusters were each isolated from the full dataset and re-clustered. We re-normalized the data subset using the same protocol as explained in section 4 in methods. The number of PCs used for UMAP dimensionality reduction was different for each cell type, 4, 8, 10, 6, 5, for Neuron, Oligodendrocytes, Microglia, Astrocytes, and OPC, respectively. We then employed Seurat’s *FindNeighbors* and *FindClusters* functions to identify unique cell states or subclusters (resolution=0.1, 0.2, 0.2, 0.05, 0.15). Additionally, we used the Garnett protocol to examine nuclei in each expression state within each cell type to detect and remove those nuclei that did not resemble a trustworthy expression profile from downstream analyses. We dropped 1 subject-specific subcluster in Neurons. After this final stage of QC, we ended with 67 of the 74 brains and 294,114 of the 1,102,459 nuclei.

### Differential proportion analysis

To identify associations between cell-type transcriptional state and disease status or genetic strata (ADAD, sAD, TREM2, TREM2_reduced, rs158276), we employed linear regression models to test each subject’s cell state compositions. More explicitly, the number of nuclei a subject had in a specific cell state was divided by the subject’s total nuclei count for that cell type creating a proportion. The proportions were normalized using a cube root transformation and were corrected

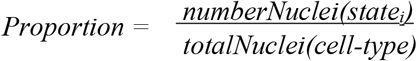

by sex, age of death (AOD), and disease status depending on the variable of interest. We removed subjects that contributed fewer than 60 nuclei to the cell type cluster. The TREM2 analyses only included the 31 sAD subjects. We utilized glm, a standard function in R, to implement the model.

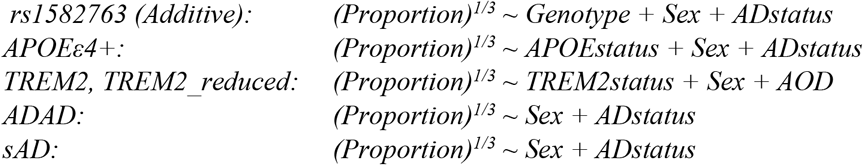

### Cell state differential expression

To determine if there was unique functionality or potentially altered cell states due to disease, we fitted a linear mixed model that predicted the expression level of each gene for the individual nuclei by cell state and corrected for the subject of origin and sex^63^. Control, sAD, and ADAD subjects were used to calculate cell state differentially expressed genes. Expression levels were extracted from the Seurat objects using *GetAssayData* with the ‘slot’ parameter set to ‘counts’. Age of death (AOD) was not included in the model because AOD is correlated with ADAD status. R package nebula version 1.1.5 was used to implement the model, including parameters for a zero-inflated negative binomial distribution and the random effect of the subject of origin^64^. The number of UMI’s per nuclei was already corrected for during *SCTransformation*, so the model did not need to account for the number of UMI’s.

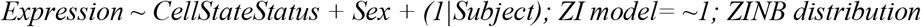

Within each cell type, differential expression (DE) was calculated on each cell state versus all other states and each state against each other state individually.

### Genetic factor differential expression

Differentially expressed genes were identified within each cell type by genetic status, namely ADAD, TREM2, or sAD vs. Controls and *APOE*ε4-vs. *APOE*ε4+. The nuclei for each status (ADAD, TREM2 variant carriers, sAD, and APOEe4+) were isolated by groups. Each group was individually compared to controls (neuropath free controls or APOEe4-) using linear mixed models as explained above. The following model was used:

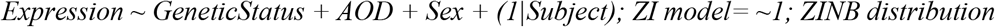

Age of Death (AOD) was not included when analyzing ADAD and *APOE*ε4+. This model was run on the entire cell type as a whole and each individual cell state within the cell types.

### Pathway analysis

The upregulated genes identified for each cell state were used in a subsequent pathway analysis. We used the R-based application Enrichr hosted by the Ma’ayan Laboratory^65,66^. We used gene sets to determine pathway enrichment by using the “KEGG 2021 Human” or “GO Biological Process 2021” gene sets. Downregulated genes were also run in the APOE-high neuron analysis.

### Microglia expression states

We collected 12 gene sets (Aging^38^, Homeostatic^67^, Lipid-droplet-accumulating^68^, Neurodegenerative^24^,Proliferative-region-associated^69^, Injury-responsive^70^, Activated-response^71^, Interferon-response^71^, Human-Alzheimer’s^25^, Disease-associated^23^, LRRK2, and Granulin microglia) associated with different microglia functional states that had been described in the literature. Each set was split into their up and down-regulated gene lists. A hypergeometric test was performed to identify which microglia transcriptional states previously reported were recapitulated in this dataset.

### Neuronal *APOE* and *MHC-I* coexpression

We followed the methods outlined by Zalocusky *et al*.^*18*^. Briefly, MAGIC^72^ was used to impute gene expression, *APOE* expression greater than two standard deviations marked *APOE*-high expression, and the genes *HLA-A, HLA-B, HLA-C, HLA-E, HLA-F*, and *B2M* were summed to represent *MHC-I* expression. Correlation between *APOE* and *MCH-I* expression was calculated using the native *cor*.*test* function in R. Genes differentially expressed between *APOE-*high and *APOE-*low neurons were identified using linear mixed models as previously described (*nebula* R package; model: expression ∼ apoeHigh + Sex). These DE analyses were performed only on the neuro.3 and neuro.5 cell states split by AD condition. Enriched pathways in the significantly upregulated genes were identified using EnrichR as previously described.

### Characterization of loci identified in AD risk GWAS

A list of genes identified through AD GWAS was collected. We started with 89 genes from 39 different loci that were previously prioritized^*13,44,45*^. We then added in all 878 genes from all the significant loci. This list was filtered by genes in our snRNA-seq data set, leaving 540 genes in 46 different loci. The significant cell state DE results were queried for the genes within our curated lists and extracted by cell type. We did not include cell states that contained less than 5% of that cell type’s nuclei. For DE analyses that were a cell state against all others (e.g. mic.1 vs all other mic cell states), we removed gene hits that had a negative estimate. We calculated the log2 fold change from the estimates provided by nebula by using this equation:

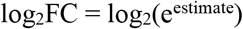

If a gene had multiple hits within a single cell type, we selected the maximum log fold change in our plots. The log2FC values greater than three were capped at three to preserve visual variability. For each gene in our gene list, we calculated the mean expression in each cell type.

### ADAD-specific microglia cluster validation

To confirm the biological existence of Mic.4, a microglia expression state composed of ADAD subjects, single microglia cells (n=1551) from 5xFAD mice were collected as described in ^73^. Protein encoding mouse genes were converted to their human orthologs using biomaRt. The data were normalized using *SCTransform*, regressing out numbers of genes and UMIs. Seurat’s best practices workflow was followed to integrate the mouse and human microglia using the human microglia as the reference dataset. Seven clusters were assigned using *FindNeighbors* and *FindClusters* with the first eight principal components and a resolution of 0.2 as input. A total of 297 mouse cells and 1413 of the 1429 human Mic.4 nuclei were recaptured in the post-integration cluster 2. The mouse cells were then re-isolated. A human Mic.4 signature score was calculated for each mouse cell by running the 412 upregulated genes that passed multiple test corrections into Seurat’s *AddModuleScore*. The significant pairwise differences in cluster scores were calculated using the linear mixed model *moduleScore ∼ cluster, ziformula=∼0, family=’gaussian’* executed using *glmmTMB*.

### TREM2-enriched microglia cluster validation

Human single nucleus data from the ROSMAP samples (DLPFC) generated as described in^16^ were used to confirm the Mic.2 cell state, enriched for TREM2 p.R47H, p.R62H and p.H157Y. The ROSMAP data has 11 sAD, 11 TREM2 R62H, and 10 control subjects with 3,986 microglia. The microglia were isolated and normalized using Seurat’s *SCTransform* function with “return.only.var.genes” set to FALSE and regressing out “nCount_RNA” and “nFeature_RNA”. The microglia were then integrated using 3000 features in *SelectIntegrationFeatures, PrepSCTintegration*, our data as a reference in *FindIntegrationAnchors*, and *IntegrateData*. To identify which nuclei fell into our original clusters, the integrated data was clustered using the first ten principal components as input for *FindNeighbors* and a resolution of 15 in *FindClusters*. This shattered the data finding 147 clusters. We assigned each of these 147 clusters an ‘original’ cluster identity by isolating our cohort of nuclei from the individual clusters and identifying the most common original ID. This ID was then transferred to the ROSMAP nuclei, similar to a nearest neighbor classifier^74^. These cluster identities were mapped to the pre-integrated normalized ROSMAP data. To measure the accuracy of our label transfer, a mic.2 signature score was calculated for each nucleus by running the mic.2 significantly upregulated genes into Seurat’s A*ddModuleScore*. Out of the 125 upregulated genes, 114 were present in the ROSMAP data. The significant pairwise differences between clusters were calculated using the linear mixed model *moduleScore ∼ cluster + AOD + Sex + (1*|*subject), ziformula=∼0, family=’gaussian’* executed using *glmmTMB*. The previously described ‘Differential Proportion Analysis’ methods were followed to verify the enrichment of TREM2 R62H nuclei in the ROSMAP Mic.2 cluster.

### TREM2-enriched oligodendrocyte cluster validation

The two oligodendrocyte clusters (29,478 nuclei) in the ROSMAP data were integrated with our oligodendrocytes and labeled with our original cluster identities as described above. A total of 264 clusters were identified using eight PCA dimensions at a resolution of 15. Because of the large number of nuclei and the reduced variability of our Oligodendrocytes, many of the Oligodendrocyte cell states shared upregulated genes. To find a clear signal for Oligo.5, we removed all upregulated genes from Oligo.5 that had a larger β (estimate) in another cell type. This left 77 of the 638 upregulated genes. 73 of those 77 were present in the ROSMAP data and used to calculate the Oligo.5 signature score. Pairwise significant differences in signature score and enrichment of TREM2 R62H variant carriers were calculated as described above. The previously described ‘Differential Compositional Analysis’ methods were followed to verify the enrichment of TREM2 R62H nuclei in the ROSMAP Oligo.5 cluster.

### Rs1582763-enriched microglia cluster validation

Using the same cluster-labelled ROSMAP microglia as described above, we calculated a mic.3 signature score. 192 of the 207 upregulated Mic.3 genes were present in the ROSMAP data. Pairwise significant differences in signature score and enrichment of rs1582763 carriers were calculated as described above. The previously described ‘Differential Compositional Analysis’ methods were followed to verify the enrichment of rs1582763 nuclei in the ROSMAP Mic.3 cluster.

### Data visualization browser

We developed the Single Nucleus Alzheimer disease RNA-seq Explorer (SNARE) to host the single nucleus expression data using the cellxgene^75^ platform. It can be publically accessed at http://ngi.pub/SNARE. We include the UMAP representations for the full data set (includes all cell types) and the individual cell type subclustering.

## Data Availability

The single nucleus data from the Knight ADRC is publicly available by request from the National Institute on Aging Genetics of Alzheimer's Disease Data Storage Site (NIAGADS) with accession number NG00108 (https://www.niagads.org/datasets/ng00108). To access the data from the DIAN brain bank, special request must be made using this URL: https://dian.wustl.edu/our-research/for-investigators/. The 5xFAD mouse microglia data are located in the Gene Expression Omnibus (GEO database) under the accession number GSE141917 (https://www.ncbi.nlm.nih.gov/geo/query/acc.cgi?acc=GSE1419177). The ROSMAP single nucleus RNA sequencing data is available at Synapse under Synapse ID syn21125841 (https://www.synapse.org/%23!Synapse:syn21125841/wiki/597278). Custom code used to analyze the snRNA-seq data and datasets generated and/or analyzed in the current study are available from the corresponding authors upon request.

## Code availability

Custom code used to analyze the snRNA-seq data and datasets generated and/or analyzed in the current study are available from the corresponding authors upon request.

## Supplementary Material

Locus_Plots – pdf file

DEG_by_cell_state – zip folder of xlsx files

## Funding

This work was possible thanks to the following governmental grants from the National institute of Health: NIA R01AG057777 (OH), R56AG067764 (OH) P30AG066444 (JCM), P01AGO26276 (JCM), U19AG032438 (RJB), R01AG044546 (CC), P01AG003991 (JCM), RF1AG053303 (CC), RF1AG058501 (CC), U01AG058922 (CC), NINDS R01NS118146 (BAB), RFNS110809 (CMK), R01AG062734 (CMK), U01AG072464 (OH, CMK), the BrightFocus Foundation (CMK), and the Chan Zuckerberg Initiative (CZI). O.H. is an Archer Foundation Research Scientist.

This work was supported by access to equipment made possible by the Hope Center for Neurological Disorders, and the Departments of Neurology and Psychiatry at Washington University School of Medicine.

## Acknowledgments

Data collection and sharing for this project was supported by The Dominantly Inherited Alzheimer Network (DIAN, U19AG032438) funded by the National Institute on Aging (NIA),the Alzheimer’s Association (SG-20-690363-DIAN), the German Center for Neurodegenerative Diseases (DZNE), Raul Carrea Institute for Neurological Research (FLENI), Partial support by the Research and Development Grants for Dementia from Japan Agency for Medical Research and Development, AMED, and the Korea Health Technology R&D Project through the Korea Health Industry Development Institute (KHIDI), Spanish Institute of Health Carlos III (ISCIII), Canadian Institutes of Health Research (CIHR), Canadian Consortium of Neurodegeneration and Aging, Brain Canada Foundation, and Fonds de Recherche du Québec – Santé. This manuscript has been reviewed by DIAN Study investigators for scientific content and consistency of data interpretation with previous DIAN Study publications. We acknowledge the altruism of the participants and their families and contributions of the DIAN research and support staff at each of the participating sites for their contributions to this study The results published here are in part based on data obtained from the AD Knowledge Portal (https://adknowledgeportal.org). Study data were provided by the Rush Alzheimer’s Disease Center, Rush University Medical Center, Chicago. Data collection was supported through funding by NIA grants P30AG10161 (ROS), R01AG15819 (ROSMAP; genomics and RNAseq), R01AG17917 (MAP), R01AG30146, R01AG36042 (5hC methylation, ATACseq), RC2AG036547 (H3K9Ac), R01AG36836 (RNAseq), R01AG48015 (monocyte RNAseq) RF1AG57473 (single nucleus RNAseq), U01AG32984 (genomic and whole exome sequencing), U01AG46152 (ROSMAP AMP-AD, targeted proteomics), U01AG46161(TMT proteomics), U01AG61356 (whole genome sequencing, targeted proteomics, ROSMAP AMP-AD), the Illinois Department of Public Health (ROSMAP), and the Translational Genomics Research Institute (genomic). Additional phenotypic data can be requested at www.radc.rush.edu.

Figure 1 was created with BioRender.com

## Competing Interests

CC receives research support from: Biogen, EISAI, Alector and Parabon. The funders of the study had no role in the collection, analysis, or interpretation of data; in the writing of the report; or in the decision to submit the paper for publication. CC is a member of the advisory board of Vivid genetics, Halia Therapeutics and ADx Healthcare.

---

**Extended Figure 1.**
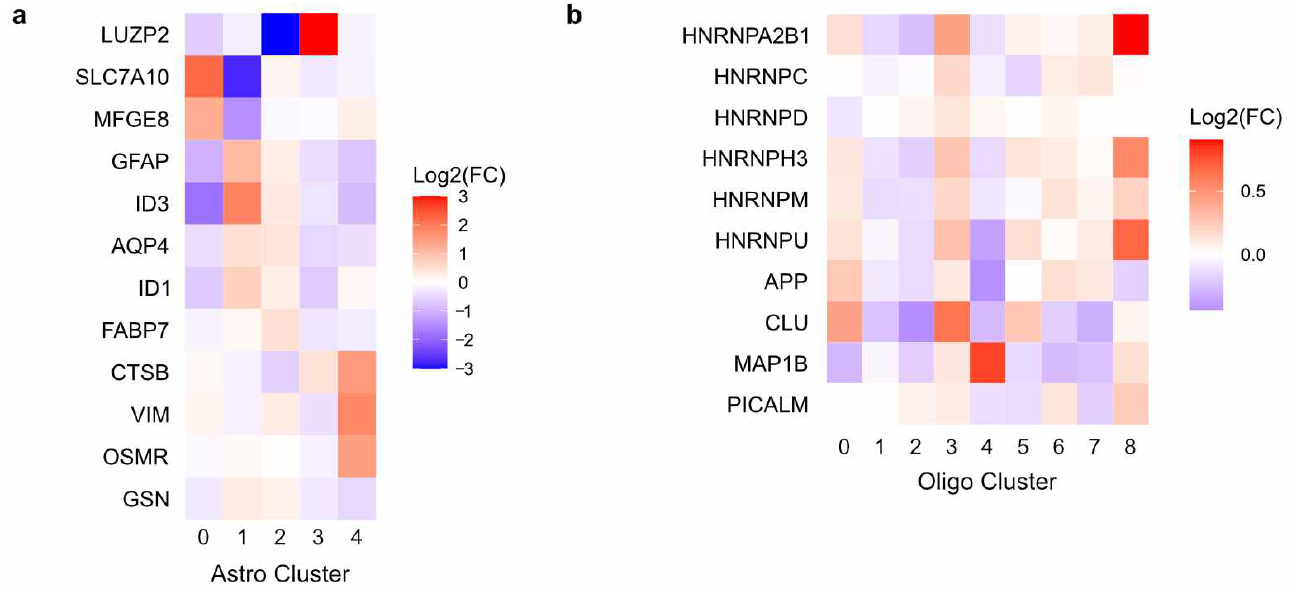
Gene expression patterns in astrocyte and oligodendrocytes clusters enriched in ADAD brains. **a)** A heatmap shows the log2 fold change (Log2FC) of astrocyte marker genes taken from Habib *et al* within the astrocyte subclusters. *LUZP2, SLC7A10*, and *MFGE8* mark GFAP-low astrocytes (resting); *GFAP, ID3, AQP4, ID1*, and *FABP7* mark GFAP-high astrocytes (activated); *CTSB, VIM, OSMR*, and *GSN* mark disease associated astrocytes (DAA). **b)** A heatmap shows the Log2FC of HNRNP genes and AD risk genes controlled by the HNRNP gene family within the oligodendrocyte subclusters.

**Extended Figure 2.**
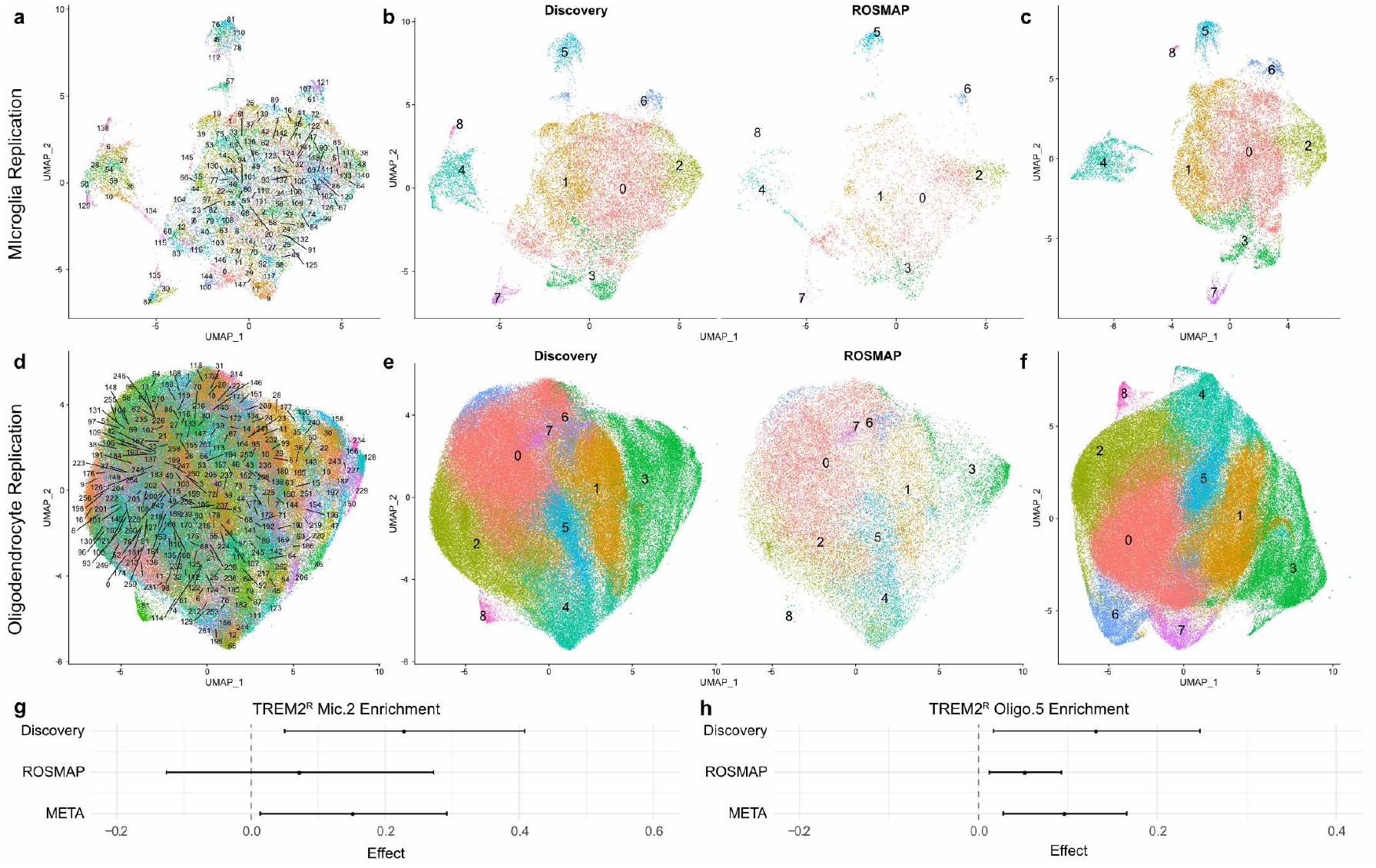
*TREM2* associated clusters replicated in ROSMAP cohort. **a**,**d)** UMAPs of discovery cohort (Knight ADRC and DIAN) and ROSMAP microglia (a) and oligodendrocytes (d) post integration. A resolution of 15 was used to identify a large number of clusters to facilitate the mapping of the discovery cohort cluster identities onto the ROSMAP nuclei. **b**,**e)** UMAPs of microglia (b) and oligodendrocytes (e) post cluster identity mapping, split by cohort. **c**,**f)** UMAPs of discovery cohort microglia (c) and oligodendrocytes (f) prior to ROSMAP integration. **g**,**h)** Tree plots of the Mic.2 (g) and Oligo.5 enrichment within TREM2 reduced activation variant carriers (TREM2^R^). The linear regression effect values are shown for the independent discovery and ROSMAP cohorts as well as the merged meta-analysis results.

## Notes

### Competing Interest Statement

Carlos Cruchaga (CC) receives research support from: Biogen, EISAI, Alector and Parabon. The funders of the study had no role in the collection, analysis, or interpretation of data; in the writing of the report; or in the decision to submit the paper for publication. CC is a member of the advisory board of Vivid genetics, Halia Therapeutics and ADx Healthcare.

### Author Declarations

The Human Research Protective Office of Washington University in St. Louis gave ethical approval for this work.

### Summary of Updates

Added author Ricardo D'Oliveira Albanus

